# Uncovering Spatiotemporal Dynamics of the Corticothalamic Network during Seizures

**DOI:** 10.1101/2023.08.21.23294382

**Authors:** Saarang Panchavati, Atsuro Daida, Benjamin Edmonds, Makoto Miyakoshi, Shingo Oana, Samuel S. Ahn, Corey Arnold, Noriko Salamon, Raman Sankar, Aria Fallah, William Speier, Hiroki Nariai

**Affiliations:** Department of Bioengineering, University of California, Los Angeles, CA, USA; Department of Radiological Sciences, University of California, Los Angeles, CA, USA; Division of Pediatric Neurology, Department of Pediatrics, UCLA Mattel Children’s Hospital, David Geffen School of Medicine, Los Angeles, CA, USA; Department of Psychiatry and Behavioral Neuroscience, University of Cincinnati, Cincinnati Children’s Hospital Medical Center, Cincinnati, OH, USA; Department of Radiology, UCLA Medical Center, David Geffen School of Medicine, Los Angeles, CA, USA; The UCLA Children’s Discovery and Innovation Institute, Los Angeles, CA, USA; Department of Neurosurgery, UCLA Medical Center, David Geffen School of Medicine, Los Angeles, CA, USA

## Abstract

**Objective:** Although the clinical efficacy of deep brain stimulation targeting the anterior nucleus (AN) and centromedian nucleus (CM) of the thalamus has been actively investigated for the treatment of medication-resistant epilepsy, few studies have investigated dynamic ictal changes in corticothalamic connectivity in human EEG recording. This study aims to establish the complex spatiotemporal dynamics of the ictal corticothalamic network associated with various seizure foci.

**Methods:** We analyzed ten patients (aged 2.7–28.1) with medication-resistant focal epilepsy who underwent stereotactic EEG evaluation with thalamic coverage. We examined both undirected and directed connectivity, incorporating coherence and spectral Granger causality analysis (GCA) between the diverse seizure foci and thalamic nuclei (AN and CM).

**Results:** In our analysis of 36 seizures, coherence between seizure onset and thalamic nuclei increased across all frequencies, especially in slower bands (delta, theta, alpha). GCA showed increased information flow from seizure onset to the thalamus across all frequency bands, but outflows from the thalamus were mainly in slower frequencies, particularly delta. In the subgroup analysis based on various seizure foci, the delta coherence showed a more pronounced increase at CM than at AN during frontal lobe seizures. Conversely, in limbic seizures, the delta coherence increase was greater at AN compared to CM.

**Interpretation:** It appears that the delta frequency plays a pivotal role in modulating the corticothalamic network during seizures. Our results underscore the significance of comprehending the spatiotemporal dynamics of the corticothalamic network during seizures, and this knowledge could guide personalized neuromodulation treatment strategies.

**Summary for Social Media:** Twitter handles: **@saarang_p; @BillSpeier**

*What is the current knowledge on the topic:* Although the clinical efficacy of deep brain stimulation targeting the anterior nucleus and centromedian nucleus of the thalamus has been actively investigated for the treatment of medication-resistant epilepsy, few studies have investigated dynamic ictal changes in corticothalamic connectivity in human EEG recording.

*What question did this study address:* This study aimed to establish the complex spatiotemporal dynamics of the ictal corticothalamic network associated with various seizure foci.

*What does this study add to our knowledge:* The delta frequency plays a pivotal role in modulating the corticothalamic network during seizures. There are seizure-onset dependent spatiotemporal dynamics of the ictal corticothalamic network.

*How might this potentially impact on the practice of neurology:* This knowledge could guide personalized neuromodulation treatment strategies.

## INTRODUCTION

Epilepsy affects approximately 1% of the population and poses a significant public health burden.^1^ While many patients with epilepsy respond well to medication, one-third of individuals develop medication resistance.^2^ Neuromodulation has emerged as a promising approach for medication-resistant epilepsy (MRE) patients who are not suitable candidates for resective surgery. Current neuromodulation options include vagus nerve stimulation (VNS),^3^ deep brain stimulation (DBS),^4^ and responsive neurostimulation (RNS).^5^

Among these approaches, thalamic stimulation has shown promise in treating MRE due to the thalamus’ role as a central hub in the cortical network. Animal and human studies using neuroimaging^6, 7^ and neurophysiology^8–10^ have provided evidence supporting the critical involvement of the thalamus in the generation, maintenance, and termination of neocortical seizures. The clinical efficacy of DBS targeting the anterior nucleus (AN) of the thalamus has been extensively studied, and studies have shown efficacy in reducing temporal and limbic seizures,^4, 11^ presumably due to the strong connection between AN and the temporal lobe/limbic system.

However, it has been found that the efficacy of AN stimulation is suboptimal in seizures other than temporal or limbic onset.^4^ As a result, some researchers have investigated the stimulation of the centromedian nucleus (CM) in the context of Lennox-Gastaut syndrome (LGS),^12^ generalized,^13, 14^ and frontal lobe epilepsy.^15^ Recent studies, including a randomized controlled trial, have demonstrated promising results for CM stimulation in patients with LGS.^16^ This suggests that the CM may exhibit stronger connections to frontal and other brain areas, thereby further supporting its potential as an effective target for neuromodulation in non-temporal/limbic epilepsy. It has also been shown that ictal EEG changes, especially in the fast frequency band, are observed at various thalamic nuclei during various seizure onsets.^17, 18^ The presence of ictal EEG changes at the thalamus may justify closed-loop neuromodulation by RNS.^19^

Despite the promise, few studies have investigated changes in corticothalamic connectivity between thalamic nuclei and different seizure foci during seizures. Neuroimaging techniques, such as functional MRI, offer good spatial resolution but lack temporal resolution, making them inadequate for elucidating dynamic temporal changes in seizure networks. On the other hand, EEG provides excellent temporal resolution, but sampling signals from the thalamus is challenging unless clinically indicated. However, several recent studies using stereotactic EEG (SEEG) demonstrated that in temporal lobe seizures, active involvement of the pulvinar nucleus,^10, 20^ and CM,^21^ along with AN was demonstrated. To enable personalized neuromodulation treatment, it is crucial to understand the distinct roles of each thalamic nucleus during seizures, taking into account their anatomically specific seizure onsets.

To address this knowledge gap, we aimed to investigate the dynamic connectivity changes within the corticothalamic network during seizures by employing undirected and directional connectivity analysis. We analyzed a unique cohort of patients with thalamic coverage during SEEG evaluation for thalamic neuromodulation. Considering the distinct anatomical regions, we explored the intricate interactions between the seizure onset zones and thalamic nuclei (AN and CM) across various seizure foci.

## MATERIALS AND METHODS

### Patient Cohort

This was a retrospective study. Patients with pediatric-onset epilepsy admitted to the UCLA Mattel Children’s Hospital and underwent a chronic SEEG implantation with electrodes inserted into the thalamus (AN and/or CM) from November 2020 to February 2023 were identified. Those patients were suspected of having MRE with focal onset and were possible candidates for thalamic neuromodulation. There were no exclusion criteria.

### Standard protocol approvals, registrations, and patient consent

The institutional review board at UCLA approved the use of human subjects and waived the need for written informed consent. All testing was deemed clinically relevant for patient care, and all retrospective EEG data used for this study were de-identified before data extraction and analysis. This study was not a clinical trial, and it was not registered in any public registry.

### Patient and seizure evaluation

The plan for SEEG placement was discussed at our multidisciplinary epilepsy surgery conference (consisting of epileptologists, neurosurgeons, radiologists, and neuropsychologists) and was based on the combination of data from seizure semiology, neurological examination, neuroimaging findings (MRI, PET, and magnetoencephalography), neuropsychological evaluation, and scalp EEG with emphasis primarily on seizure onset zones. Ipsilateral (to the presumed site of the presumed seizure onset) AN and CM thalamic SEEG electrodes were placed to determine whether an ictal pattern can be detected in the thalamus and provide a potential target for neuromodulation. If the patient had bilateral seizure onsets during the phase 1 monitoring, thalamic SEEG electrodes were placed on the side of the most significant seizure burden, or if unclear bilateral thalamic electrodes were placed. To enable a group analysis, for each patient, we identified up to three habitual seizures with the same cortical onset or clinical semiology, which had no preceding seizures within 30 minutes. For example, if a patient had two different clinical seizure semiologies or two seizures with similar clinical semiology but clearly different electrographic onsets, these were considered different seizure types, and six seizures were reviewed (see details in **Table 1**).

**Table 1.**
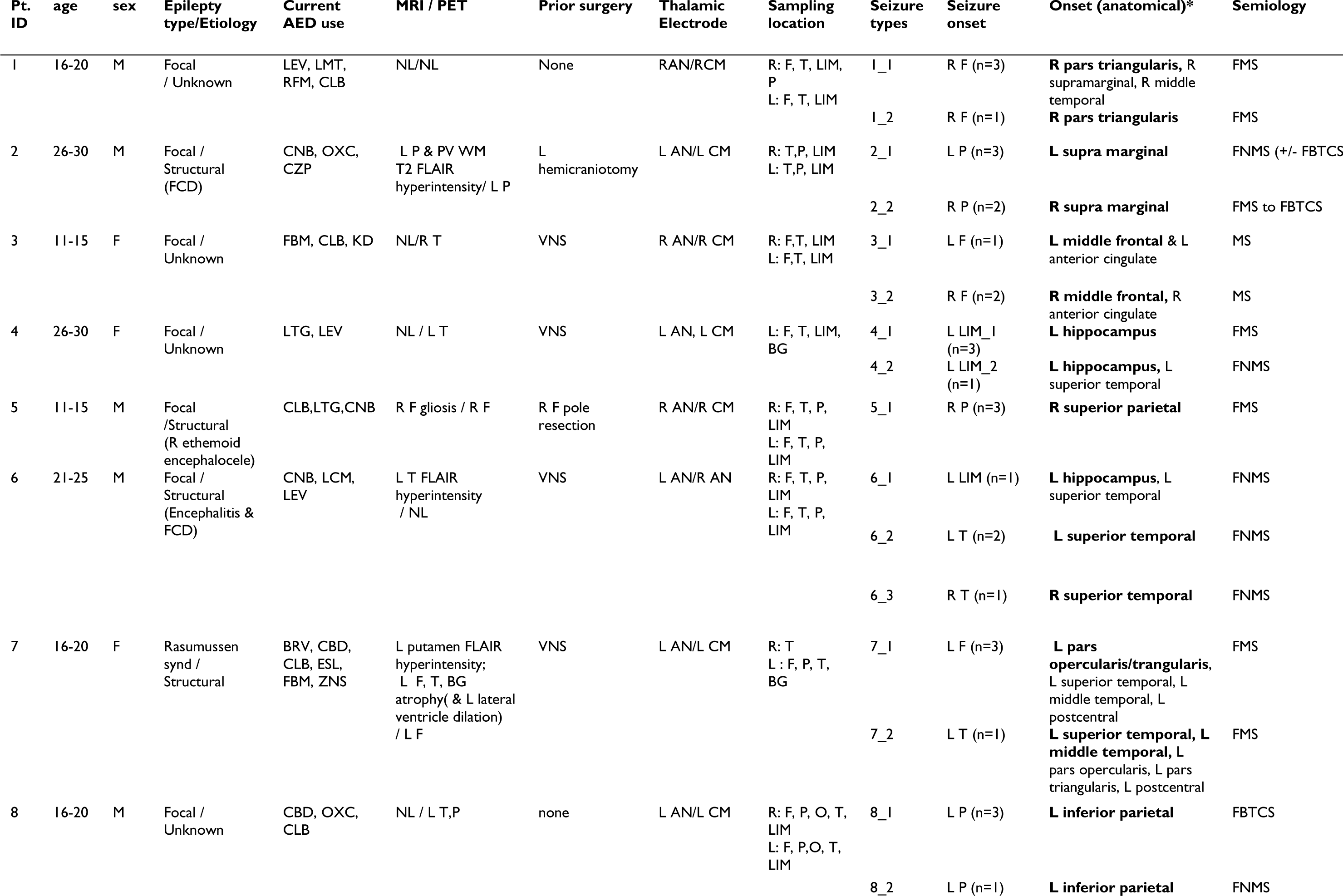
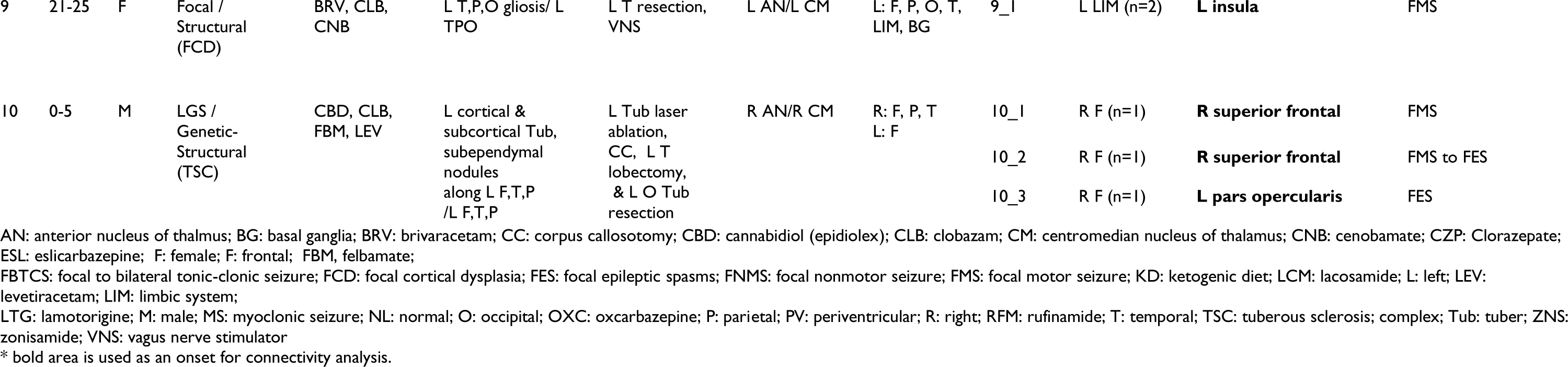
Patient Characteristics.

| Pt. ID | age | sex | Epilepsy type/Etiology | Current AED use | MRI / PET | Prior surgery | Thalamic Electrode | Sampling location | Seizure types | Seizure onset | Onset (anatomical)* | Semiology |
| --- | --- | --- | --- | --- | --- | --- | --- | --- | --- | --- | --- | --- |
| 1 | 16-20 | M | Focal / Unknown | LEV, LMT, RFM, CLB | NL/NL | None | RAN/RCM | R: F, T, LIM, P<br>L: F, T, LIM | 1_1 | R F (n=3) | **R pars triangularis**, R supramarginal, R middle temporal | FMS |
|  |  |  |  |  |  |  |  |  | 1_2 | R F (n=1) | **R pars triangularis** | FMS |
| 2 | 26-30 | M | Focal / Structural (FCD) | CNB, OXC, CZP | L P & PV WM T2 FLAIR hyperintensity/ L P | L hemicraniotomy | L AN/L CM | R: T,P, LIM<br>L: T,P, LIM | 2_1 | L P (n=3) | **L supra marginal** | FNMS (+/- FBTCs) |
|  |  |  |  |  |  |  |  |  | 2_2 | R P (n=2) | **R supra marginal** | FMS to FBTCs |
| 3 | 11-15 | F | Focal / Unknown | FBM, CLB, KD | NL/R T | VNS | R AN/R CM | R: F,T, LIM<br>L: F,T, LIM | 3_1 | L F (n=1) | **L middle frontal & L anterior cingulate** | MS |
|  |  |  |  |  |  |  |  |  | 3_2 | R F (n=2) | **R middle frontal**, R anterior cingulate | MS |
| 4 | 26-30 | F | Focal / Unknown | LTG, LEV | NL / L T | VNS | L AN, L CM | L: F, T, LIM, BG | 4_1 | L LIM_1 (n=3) | **L hippocampus** | FMS |
|  |  |  |  |  |  |  |  |  | 4_2 | L LIM_2 (n=1) | **L hippocampus**, L superior temporal | FNMS |
| 5 | 11-15 | M | Focal /Structural (R ethmoid encephalocele) | CLB,LTG,CNB | R F gliosis / R F | R F pole resection | R AN/R CM | R: F, T, P, LIM<br>L: F, T, P, LIM | 5_1 | R P (n=3) | **R superior parietal** | FMS |
| 6 | 21-25 | M | Focal / Structural (Encephalitis & FCD) | CNB, LCM, LEV | L T FLAIR hyperintensity / NL | VNS | L AN/R AN | R: F, T, P, LIM<br>L: F, T, P, LIM | 6_1 | L LIM (n=1) | **L hippocampus**, L superior temporal | FNMS |
|  |  |  |  |  |  |  |  |  | 6_2 | L T (n=2) | **L superior temporal** | FNMS |
|  |  |  |  |  |  |  |  |  | 6_3 | R T (n=1) | **R superior temporal** | FNMS |
| 7 | 16-20 | F | Rasmussen synd / Structural | BRV, CBD, CLB, ESL, FBM, ZNS | L putamen FLAIR hyperintensity; L F, T, BG atrophy( & L lateral ventricle dilation) / L F | VNS | L AN/L CM | R: T<br>L: F, P, T, BG | 7_1 | L F (n=3) | **L pars opercularis/trangularis**, L superior temporal, L middle temporal, L postcentral | FMS |
|  |  |  |  |  |  |  |  |  | 7_2 | L T (n=1) | **L superior temporal, L middle temporal, L pars opercularis, L pars triangularis, L postcentral** | FMS |
| 8 | 16-20 | M | Focal / Unknown | CBD, OXC, CLB | NL / L T,P | none | L AN/L CM | R: F, P, O, T, LIM<br>L: F, P,O, T, LIM | 8_1 | L P (n=3) | **L inferior parietal** | FBTCs |
|  |  |  |  |  |  |  |  |  | 8_2 | L P (n=1) | **L inferior parietal** | FNMS |
| 9 | 21-25 | F | Focal /<br>Structural<br>(FCD) | BRV, CLB,<br>CNB | L T,P,O gliosis/ L<br>TPO | L T resection,<br>VNS | L AN/L CM | L: F, P, O, T,<br>LIM, BG | 9_1 | L LIM (n=2) | **L insula** | FMS |
| 10 | 0-5 | M | LGS /<br>Genetic-Structural<br>(TSC) | CBD, CLB,<br>FBM, LEV | L cortical &<br>subcortical Tub,<br>subependymal<br>nodules<br>along L F,T,P<br>/L F,T,P | L Tub laser<br>ablation,<br>CC, L T<br>lobectomy,<br>& L O Tub<br>resection | R AN/R CM | R: F, P, T<br>L: F | 10_1 | R F (n=1) | **R superior frontal** | FMS |
|  |  |  |  |  |  |  |  |  | 10_2 | R F (n=1) | **R superior frontal** | FMS to FES |
|  |  |  |  |  |  |  |  |  | 10_3 | R F (n=1) | **L pars opercularis** | FES |

AN: anterior nucleus of thalamus; BG: basal ganglia; BRV: brivaracetam; CC: corpus callosotomy; CBD: cannabidiol (epidiolex); CLB: clobazam; CM: centromedian nucleus of thalamus; CNB: cenobamate; CZP: Clorazepate; ESL: eslicarbazepine; F: female; F: frontal; FBM, felbamate;
FBTCS: focal to bilateral tonic-clonic seizure; FCD: focal cortical dysplasia; FES: focal epileptic spasms; FNMS: focal nonmotor seizure; FMS: focal motor seizure; KD: ketogenic diet; LCM: lacosamide; L: left; LEV: levetiracetam; LIM: limbic system;
LTG: lamotrigine; M: male; MS: myoclonic seizure; NL: normal; O: occipital; OXC: oxcarbazepine; P: parietal; PV: periventricular; R: right; RFM: rufinamide; T: temporal; TSC: tuberous sclerosis; complex; Tub: tuber; ZNS: zonisamide; VNS: vagus nerve stimulator
\* bold area is used as an onset for connectivity analysis.

### SEEG placement and confirmation

BrainLab Elements software was used for planning the electrodes to the intended targets using T1-weighted sequences, and the trajectories were guided by a gadolinium-enhanced T1-weighted MRI. Target subcortical structures (including AN and CM) were identified and outlined by an experienced neuroradiologist (NS) on a case-by-case basis prior to the trajectory planning. The targets and trajectories that were planned using the MRI MPRAGE2 sequence were then co-registered to a volumetric computed tomography (CT) scan acquired after placing the patient’s head into the Leksell frame. Each electrode was placed using the Leksell coordinates obtained from the BrainLab elements software. Four contact Spencer Depth Electrodes with 2.5 mm spacing were used exclusively for thalamic targets. An intraoperative or immediate postoperative CT scan was used to rule out intracranial hemorrhage and confirm the final position and trajectory of each electrode placed. The location of electrode placement was verified post-operatively with CT co-registered with the pre-op T1-weighted MRI using the BrainLab Elements software. Outlines for target subcortical nuclei were identified and overlayed onto the postoperative CT scan. Electrode contact placement in relation to the target nuclei was then determined. For contacts placed outside of the desired nucleus, distance-to-target measurements were then recorded from the edge of the respective contact to the edge of the nucleus of interest. Our prior study showed that more than 90% of our cases had active contact in or within 1 mm of the nucleus it was intended for.^18^ For those leads with no electrode in the intended nucleus, the mean distance from the edge of the electrode to the edge of the thalamic nucleus was 0.4 mm for CM and 1.6 mm for AN. There were no complications, and specifically no intracranial hemorrhage, as a result of placing SEEG electrodes, including thalamic SEEG electrodes.

### EEG data acquisition

Intracranial EEG (iEEG) recording was obtained using Nihon Kohden (Irvine, California, USA) with a digital sampling frequency of 200 Hz or 2000 Hz and a proprietary band-pass frequency of 0.08-300 Hz. All iEEGs were part of the clinical EEG recording. The iEEG data were preprocessed using a notch filter at 60 Hz intervals to remove power-line noise. All data were downsampled to 200 Hz to ensure the same sampling rate across EEG recordings and improve computational tractability. We time-locked each seizure recording to the seizure onset and used 20 minutes of data before and after the seizure.

### EEG channel coregistration and channel selection for analysis

We obtained preoperative high-resolution 3D magnetization-prepared rapid acquisition with gradient echo (MPRAGE) T1-weighted image of the entire head. A FreeSurfer-based 3D surface image was created with the location of electrodes directly defined within the brain structure using post-implant CT images using Brainstorm software (**Figure 1**).^22, 23^ We restricted our analysis to electrodes located only in relevant regions and removed any electrodes primarily located in white matter, within abnormal structural lesions (such as tubers), outside the brain, or exhibited significant recording artifacts. A maximum of three seizure onset electrodes were defined as those immediately surrounding the clinically labeled seizure onset electrodes based on visual analysis by two board-certified clinical neurophysiologists.^18^ The analyses included all the thalamic channels (AN and CM). The subsequent quantitative analyses used the bipolar montage. For further subgroup analysis, all electrodes were categorized into seizure onset, AN, CM, frontal, temporal, parietal, occipital, basal ganglia, and limbic system based on the reconstruction using FreeSurfer-based Desikan-Killiany atlas. The limbic system electrodes included those from the hippocampus, parahippocampus, amygdala, cingulate, and insula. Then all the seizures from each category were collected and analyzed.

**Figure 1.**
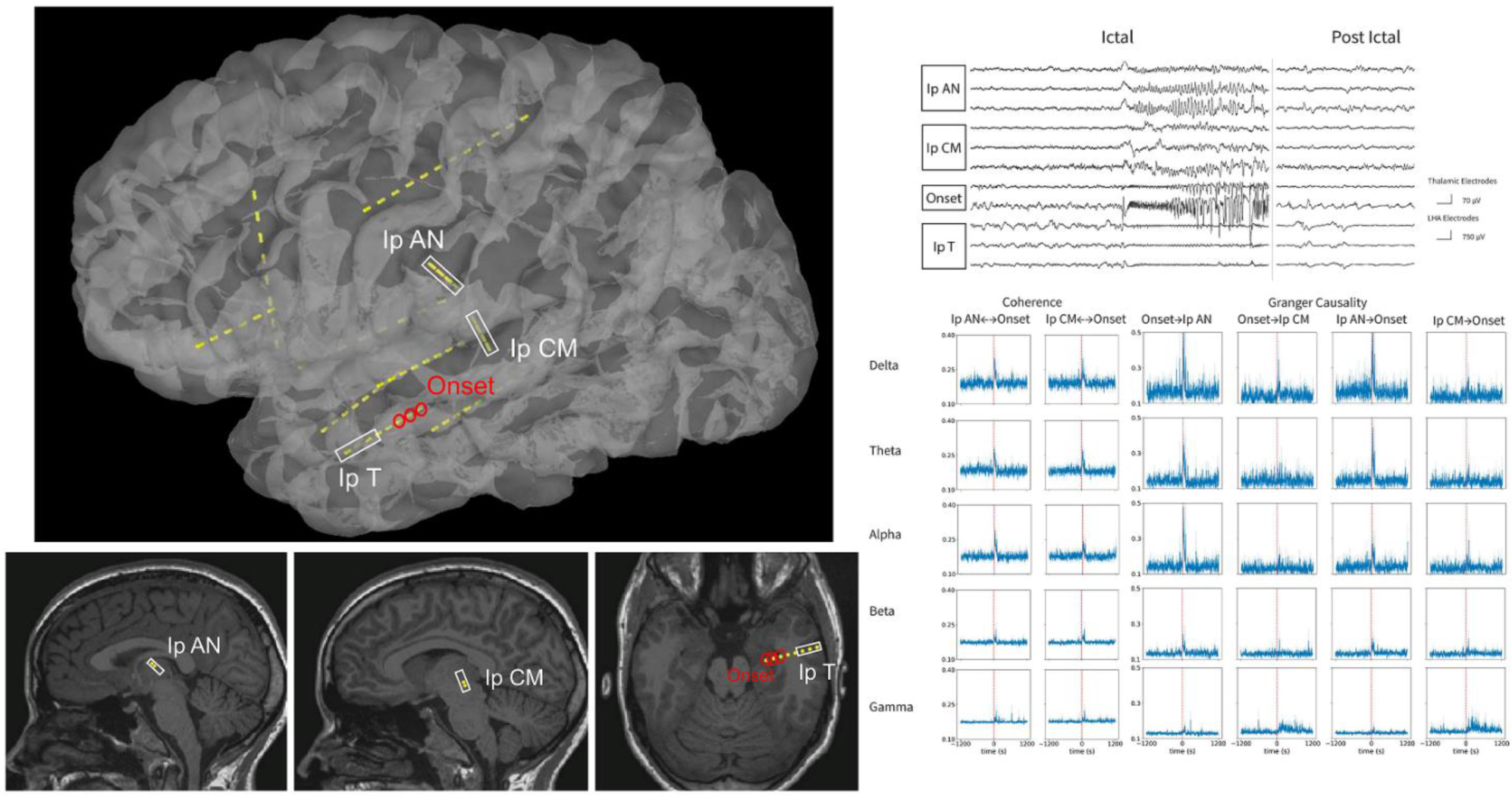
An example of SEEG placement, ictal iEEG signature, Coherence, and spectral Granger Causality for a subject with left limbic seizure. (Left) A lateral view of a 3D reconstruction of the SEEG implantation is shown at the top panel. The trajectory of the electrodes was superimposed on the sagittal view for thalamic channels and the axial view for temporal cortical and onset channels at the bottom panels. (Right) The seizure onset in the iEEG tracing is clearly observed in both thalamic and non-thalamic channels as low-voltage fast patterns at the top panel. The time courses for coherence and spectral Granger causality are presented from 20 minutes before the seizure to 20 minutes after at the bottom panels. There was a significant increase in bidirectional connectivity at the time of seizure onset for both seizures. Notably, the changes in Granger causality were inversely proportional to the frequency band, with the delta band exhibiting a more significant increase compared to the beta/gamma bands. Ip: ipsilateral; AN: anterior nucleus; CM: centromedian; T: temporal.

**Figure 2.**
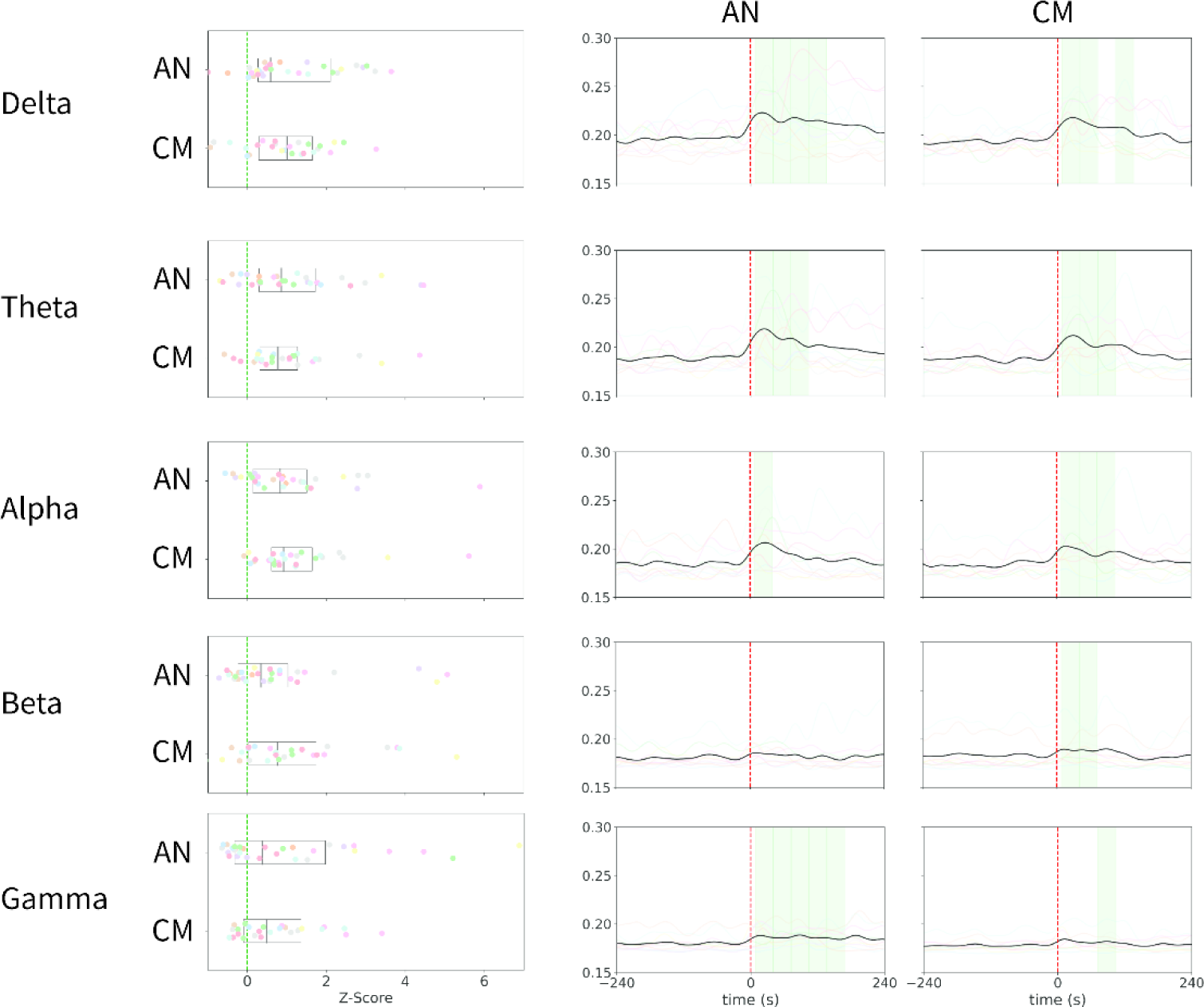
Summary of coherence analysis across seizures. The left panels illustrate the z-score difference between ipsilateral AN and onset, as well as ipsilateral CM and onset, during the 0-20s interval after the seizure compared to the baseline (20-10 mins before the seizure) for five frequency bands. Each colored dot represents an individual subject. Overall, coherence demonstrates an increase in all frequency bands during seizures. The right panels display a group analysis of coherence time courses from 4 minutes before seizure to 4 minutes after seizure for different frequency bands and interactions between AN and onset and CM and onset. The shaded green region indicates a statistically significant (Mann-Whitney U Test) increase in coherence relative to the aforementioned baseline. The calculations were performed using a sliding window of size 32s and a 4s step. The colored lines represent the average coherence across seizures for each patient, while the bolded black line represents the mean time course across all patients and seizures. There is a notable increase in connectivity between both thalamic nuclei and the seizure onset at all frequency bands except for the Beta band at AN. AN: anterior nucleus; CM: centromedian

### Connectivity Analysis

Using a bipolar montage, we segmented the SEEG data into four-second non-overlapping windows and calculated measures of connectivity on each window. This study employed coherence and spectral Granger causality to explore connectivity at the baseline (20-10 mins before the seizure) and ictal segment (0-20s after visually marked EEG seizure onset). Such connectivity values were contrasted between the baseline and the ictal segment to determine the connectivity change. We quantified connectivity between the seizure onset zones and AN and CM. We also explored connections between different brain lobes for different seizure foci. We also investigated the connectivity changes across various frequency bands: delta (1.5-4 Hz), theta (4-8 Hz), alpha (8-12 Hz), beta (12-30 Hz), and gamma (30-100 Hz) (a case example presented in **Figure 1**). The MNE-Python package^24^ was used for all preprocessing of the data.

#### Coherence

Coherence is an undirected measure of synchronization and correlation between two-time series signals at different frequencies. Values of coherence range between 0 and 1, where 1 represents the highest degree of synchrony between two signals. Coherence has widely been used in EEG data analysis to understand functional connectivity between different brain regions.^25^ We calculated pairwise coherence between electrodes using the MNE-connectivity open-source Python package.^24^ We averaged coherence values within each of the frequency bands of interest. Subsequently, these values were averaged across the different electrode interactions. For example, coherence between electrodes in the thalamus and at seizure onset was averaged to obtain a coherence value for thalamic-onset interactions at a particular time window and frequency band.

#### Granger Causality

In the time domain, Granger causality analysis (GCA) is a statistical measure of how one time series influences another based on two autoregressive models. If the inclusion of the history of time series A with the history of time series B reduces the prediction error for B compared to just the history of B, then A is said to Granger cause time series B. While this is a directed measure, causality can be bidirectional, where two signals can influence each other. Like coherence, granger causality has been a common tool in neural signal analysis to capture directed functional connectivity.^26^ In this study, we used spectral GCA, which is a variation of the traditional time series GCA. Spectral Granger causality measures the fraction of the total power of a signal at a frequency contributed by another signal.^27^ We calculated spectral GCA in a similar manner to coherence, in which our final result was a value between 0 and 1 for a directed interaction (thalamus granger causes seizure onset region or vice versa) at a particular time window and frequency band. Spectral GCA was calculated using the open-source spectral-connectivity Python package (https://github.com/Eden-Kramer-Lab/spectral_connectivity/).

### Statistical Analysis

The Mann-Whitney U test was used to characterize ictal and post-ictal changes in connectivity quantitatively. We used a one-sided test to determine if there was a statistically significant increase in connectivity. The baseline was set to the connectivity (coherence values or spectral GCA values) 20 to 10 minutes before seizure onset. We then calculated the statistical significance between a 32-second sliding window with a 4-second step size and the baseline. Bonferroni correction was used to correct for multiple comparisons by adjusting the statistical significance threshold alpha (0.05) by the number of comparisons. The total number of comparisons was defined as the product of the number of windows, frequency bands, and interactions. We defined the latency of interaction as the first time at which a statistically significant increase from baseline was observed, while the duration refers to the length of time during which the increase remained statistically significant. We also conducted two-sided Mann-Whitney U tests to identify statistically significant differences between frequency bands or thalamic nuclei. In the post-hoc analysis, we used a paired Wilcoxon signed-rank test to obtain paired differences in coherence connectivity between the thalamic nuclei.

## RESULTS

### Cohort and seizure characteristics

Ten patients (five females) were identified for this study. The median age at thalamic recording was 18.4 years (range 2.7–28.1 years). An average of 87 (+/− 35.7) depth electrode contacts were placed, providing unilateral SEEG coverage in the AN/CM implant for nine patients, while one patient received bilateral AN coverage. Epilepsy etiology included tuberous sclerosis complex, Rasmussen’s encephalitis, focal cortical dysplasia (FCD), encephalocele, and unknown. Seizure types included focal motor, focal impaired awareness, focal motor to bilateral tonic-clonic, myoclonic, epileptic spasms, and startle-induced. There were no patients excluded. A total of 36 seizures were analyzed for further group analysis (the median number was 4, with a range of 2-5 per patient). (see details in **Table 1**).

### Undirected connectivity changes based on coherence analysis

Based on the coherence analysis, we demonstrated that there was a significant increase in the undirected connectivity during seizures in all frequency bands (delta, theta, alpha, beta, and gamma) at the group level, except beta band in AN (**Table 2A**). The increase in coherence change (% change) was greater in the delta band compared to the gamma band, with values of 16.2% vs. 4.8% (AN) and 14.9% vs. 5.4% (CM), respectively (p < 0.001 and p < 0.005). Also, the increase in coherence change (% change) was greater in the ipsilateral AN compared to the contralateral AN (16.2% vs. 5.9%; p = 0.03) (**Table 2A**). With the limited number of seizures contralateral to CM sampling (n = 4), we were unable to compare the coherence changes between the ipsilateral and contralateral CM.

**Table 2A:** Summary of coherence analysis.

|  |  | Onset <-> Ip_AN | Onset <-> Ip_CM | Onset <-> Co_AN |
| --- | --- | --- | --- | --- |
| **Delta** | Raw Coherence | 0.23 | 0.22 | 0.20 |
|  | %Change | 16.19% | 14.92% | 5.92% |
|  | Duration (s) | 104 | 64 | NA |
|  | Latency (s) | 3 | 7 | NA |
| **Theta** | Raw Coherence | 0.22 | 0.21 | 0.20 |
|  | %Change | 15.32% | 13.52% | 4.90% |
|  | Duration (s) | 92 | 68 | NA |
|  | Latency (s) | 3 | 7 | NA |
| **Alpha** | Raw Coherence | 0.21 | 0.21 | 0.20 |
|  | %Change | 12.00% | 16.27% | 7.09% |
|  | Duration (s) | 48 | 60 | NA |
|  | Latency (s) | 7 | -1 | NA |
| **Beta** | Raw Coherence | 0.19 | 0.19 | 0.20 |
|  | %Change | 4.94% | 8.25% | 5.00% |
|  | Duration (s) | NA | 20 | NA |
|  | Latency (s) | NA | 7 | NA |
| **Gamma** | Raw Coherence | 0.19 | 0.19 | 0.18 |
|  | %Change | 4.80% | 5.40% | -0.07% |
|  | Duration (s) | 124 | 4 | NA |

NA: Not applicable (the value could not be determined because the calculation did not reach statistical significance)
Ip: ipsilateral; Co: contralateral; AN: anterior nucleus; CM: centromedian nucleus

### Directional connectivity changes based on Granger causality analysis

Based on the Granger causality analysis, we demonstrated a significant increase in the bidirectional connectivity during seizures, except outflow (to the onset from the thalamus) in the alpha and beta bands (**Figure 3**). While the inflow of information increased during seizures across the frequency bands, the outflow of information was most prominently observed in the delta band. Also, the onset latency of the increase in functional connectivity in both directions during seizures was earlier in the slow frequencies (delta and theta) than in the fast frequencies (beta and gamma) (**Figure 4** and **Table 2B**).

**Figure 3.**
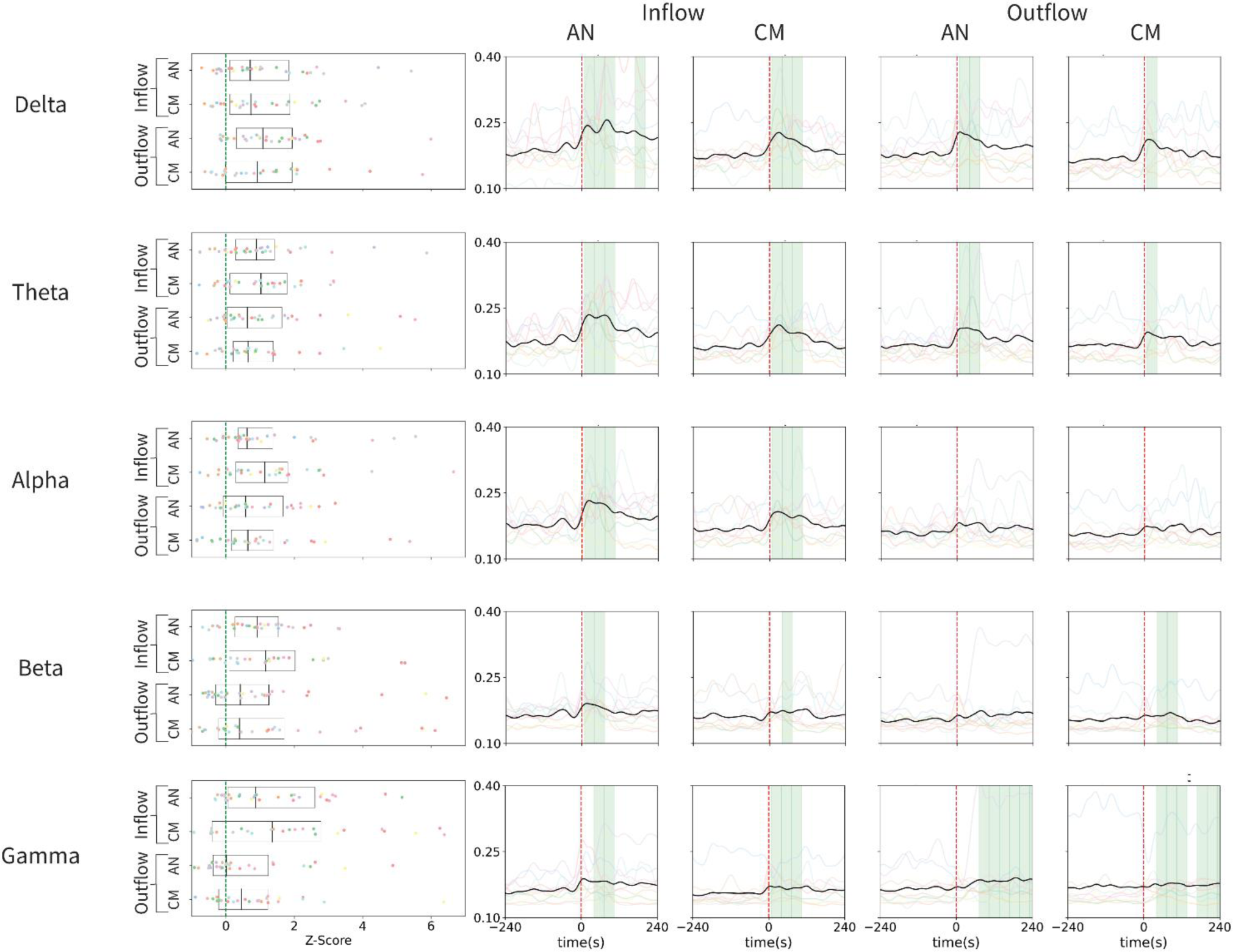
Summary of Granger causality analysis across seizures. The left panels illustrate the z-scored change in spectral Granger causality from the baseline period (20-10 mins before seizure) to the seizure window (0-10s after seizure), while the right panels depict the group analysis of the time course of spectral Granger causality from 4 minutes before seizure to 4 minutes after seizure for different electrode interactions. Inflow refers to the directed spectral Granger causality from the onset to AN or CM, while outflow refers to the directed causality from AN or CM to the onset. The shaded green regions indicate if a window exhibits a statistically significant increase in spectral Granger causality. The differently colored lines represent the mean values of all seizures for a specific patient, and the bolded black line represents the mean across all seizures. Overall, there is a clear and significant increase in bidirectional spectral Granger causality between the thalamus and cortical onset in the Delta, Theta, Beta, and Gamma frequency bands. In the Alpha band, there are only statistically significant increases in the inflow to the thalamus. AN: anterior nucleus; CM: centromedian

**Figure 4.**
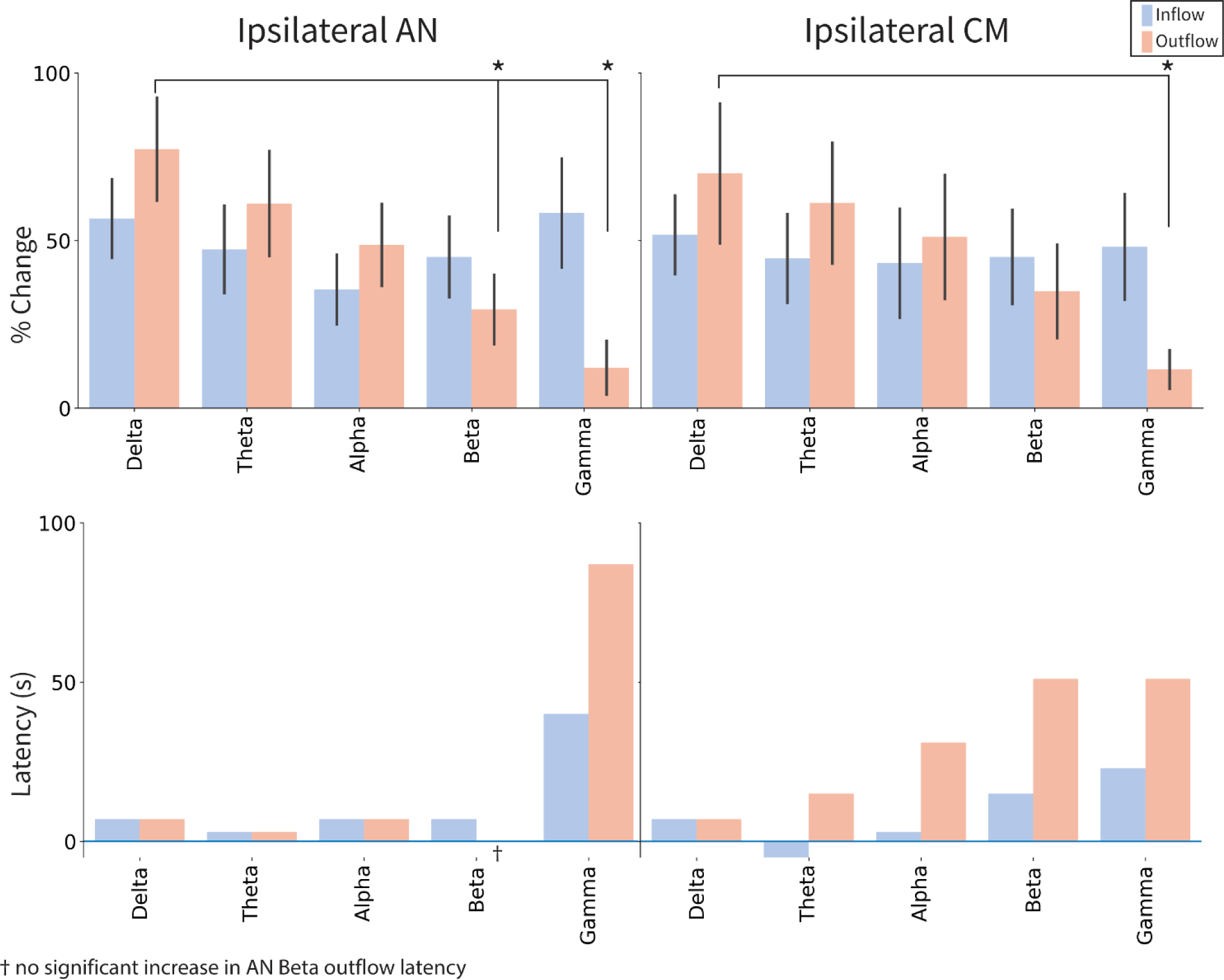
Summary of percent change and latency in Granger causality. The top panel provides a summary of the percentage change in inflow and outflow to the ipsilateral AN and CM. There was a statistically significant difference (Mann-Whitney U Test) between Delta outflow and Beta/Gamma outflow in the AN and between Delta and Gamma outflow in the CM. The bottom panel summarizes the latency, which indicates how soon after seizure onset, a significant increase occurs for ipsilateral AN and CM inflow and outflow. We observed that the latency increased as the frequency bands became faster. Of note, there was no statistically significant increase in outflow from the AN in the beta band. AN: anterior nucleus; CM: centromedian

**Table 2B:** Summary of Granger causality analysis.

|  |  | Inflow |  | Outflow |  |
| --- | --- | --- | --- | --- | --- |
|  |  | Onset -> Ip_AN | Onset -> Ip_CM | Ip_AN -> Onset | Ip_CM -> Onset |
| **Delta** | Raw GCA | 0.28 | 0.23 | 0.31 | 0.27 |
|  | %Change | 54.49% | 46.73% | 78.25% | 64.59% |
|  | Duration (s) | 40 | 92 | 80 | 40 |
|  | Latency (s) | 7 | 7 | 7 | 7 |
| **Theta** | Raw GCA | 0.25 | 0.22 | 0.26 | 0.25 |
|  | %Change | 49.81% | 42.18% | 63.01% | 55.82% |
|  | Duration (s) | 108 | 116 | 40 | 24 |
|  | Latency (s) | 3 | -5 | 3 | 15 |
| **Alpha** | Raw GCA | 0.23 | 0.22 | 0.23 | 0.22 |
|  | %Change | 35.50% | 42.02% | 47.85% | 47.76% |
|  | Duration (s) | 96 | 108 | 4 | 4 |
|  | Latency (s) | 7 | 3 | 7 | 31 |
| **Beta** | Raw GCA | 0.23 | 0.21 | 0.19 | 0.19 |
|  | %Change | 45.49% | 41.28% | 30.16% | 31.80% |
|  | Duration (s) | 20 | 4 | NA | 48 |
|  | Latency (s) | 7 | 15 | NA | 51 |
| **Gamma** | Raw GCA | 0.26 | 0.21 | 0.18 | 0.17 |
|  | %Change | 58.13% | 44.48% | 12.32% | 10.81% |
|  | Duration (s) | 55 | 40 | 8 | 92 |
|  | Latency (s) | 40 | 23 | 87 | 51 |

NA: Not applicable (the value could not be determined because the calculation did not reach statistical significance)
GCA: Granger causality analysis; Ip: ipsilateral; Co: contralateral; AN: anterior nucleus; CM: centromedian nucleus

### Connectivity changes based on seizure onset focus

We investigated undirected and directional connectivity changes based on different seizure onsets as the post-hoc analysis (all seizures were analyzed for limbic seizures). (**Figure 5**). The delta coherence showed a more pronounced increase at CM than at AN during frontal lobe seizures (14.7% vs. 8.3%; p < 0.01). Conversely, in limbic seizures, the increase was greater at AN compared to CM (14.7% vs. 7.1%; p = 0.02). In the case of parietal lobe seizures, no difference was noted between CM vs. AN (28.8% vs. 23.1%; p = 0.84) (**Figure 5**). We did not make any comparisons in the temporal neocortical seizures since the number was limited (n = 4). With the directional connectivity analysis, we demonstrated that the inflow to the thalamus from the onsets increased mostly ipsilaterally during seizures across the frequency bands but more prominently in slower frequencies (delta, theta, and alpha). Similarly, each brain region’s outflow to the thalamus was relatively limited to ipsilateral and slower frequency bands (delta, theta, and alpha) (**Supplementary Figures 1 and 2**).

**Figure 5.**
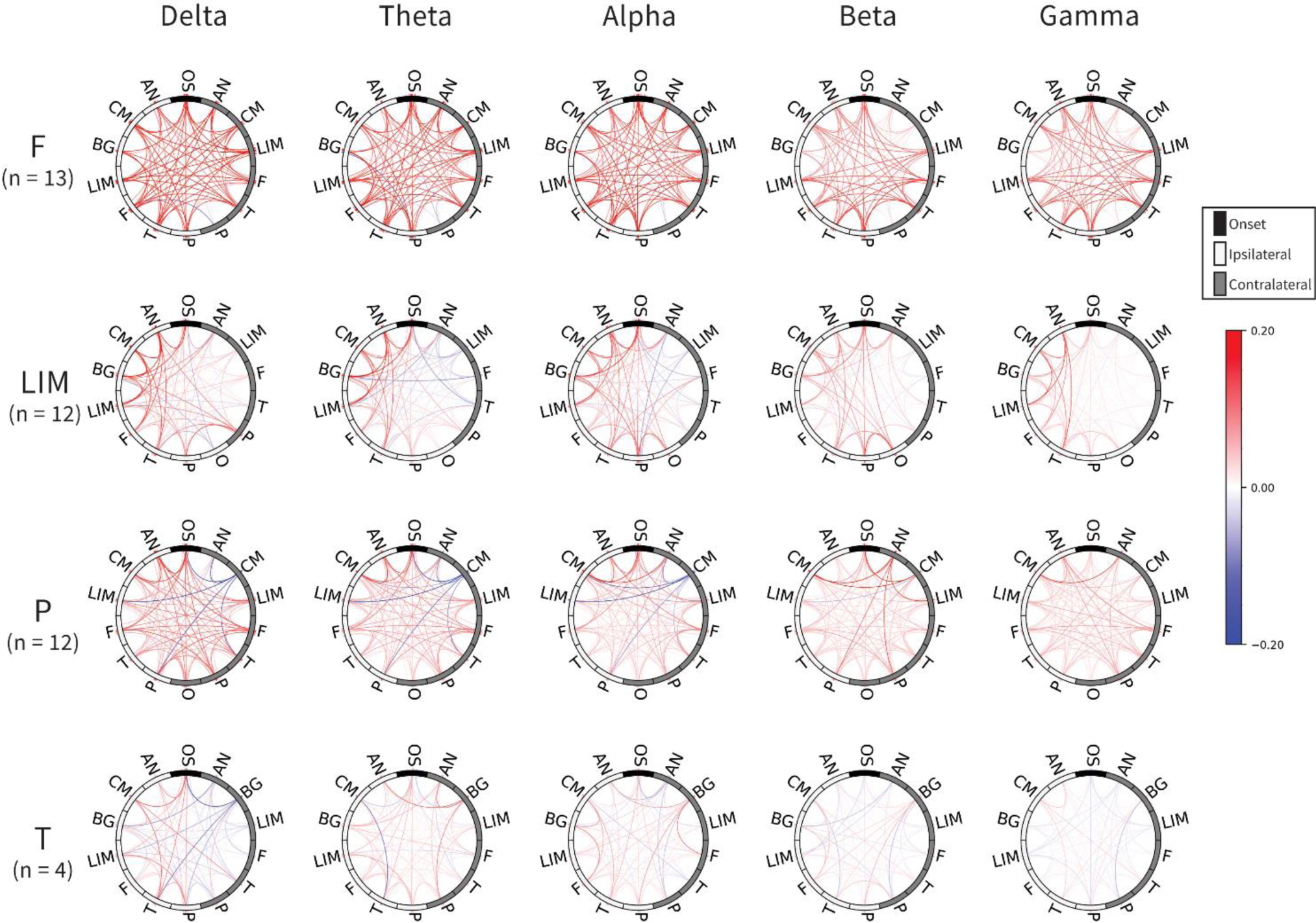
Coherence changes across different seizure foci and frequency bands. Each node on the circles represents a collection of electrodes within a specific brain region. Nodes colored white indicate ipsilateral locations to the seizure, while grey-colored nodes represent contralateral regions. A line connecting two nodes signifies a non-zero change in coherence, and the hue of the line denotes the percentage change in coherence between the aforementioned baseline and the 0-20s interval after the seizure. Seizures are categorized based on their onset brain regions. In frontal lobe seizures, there is a pronounced bilateral increase in coherence between the cortical onset and other brain regions. In limbic seizures, coherence increases are predominantly observed ipsilateral to the seizure onset. Parietal lobe seizures exhibit similar coherence patterns as frontal onset seizures but with fewer coherence increases following seizure onset. It is important to note that this figure includes the post-hoc analysis (all seizures were analyzed for limbic seizures). F: Frontal; LIM: Limbic; P: Parietal; T: Temporal; AN:Anterior Nucleus; CM: Centromedian; BG: Basal Ganglia; O: Occipital; OS: Onset; Ip: ipsilateral; Co: contralateral

## DISCUSSION

This study investigated a unique cohort of ten patients with MRE who had thalamic iEEG coverage within AN and CM. Using undirected and directed connectivity analysis, we elucidated the dynamic changes in corticothalamic connectivity between thalamic nuclei and different seizure foci during seizures. We found that coherence between seizure onset and thalamic nuclei significantly increased across all frequency bands, especially in slower frequencies (delta, theta, and alpha) compared to faster frequencies (beta and gamma). In the spectral GCA, we observed an increased information flow from seizure onset regions to the thalamus in all frequency bands. However, the outflows from the thalamus to other brain regions were primarily in slower frequency bands, particularly in the delta band. In the delta frequency range, rather than faster frequencies, the increase in directed connectivity occurred simultaneously in both inflows and outflows at the start of the ictal EEG. Our findings are consistent with the prior studies. In a study using an animal model, bidirectional cortical and thalamic interactions during absence seizures were demonstrated.^8^ Another study showed hippocampal seizures suppressed the intralaminar thalamic and brainstem arousal system, inhibiting the cortical function (network inhibition hypothesis).^28^ Our results provide compelling evidence that the delta frequency serves as a distinct marker of inhibitory outflow from the thalamus to the seizure onset. This finding strongly aligns with the traditional theory proposing that the delta component observed in the 3 Hz spike-wave discharges during absence seizures originate from the inhibitory corticothalamic input.^29^

Our results underscore the significance of comprehending the spatiotemporal dynamics of the corticothalamic network during seizures. We observed that during frontal lobe seizures, the increase in delta coherence was more prominent in CM compared to AN. Conversely, in limbic seizures, the delta coherence increase was greater in AN than in CM. This finding aligns with previous studies utilizing noninvasive structural and functional imaging techniques, which have consistently demonstrated strong physiological thalamocortical connections between the frontal lobe and CM, as well as between the limbic area and AN.^30^ Our findings are in line with the results of a recent SEEG study that investigated changes in ictal connectivity during temporal seizures. The study reported increased connectivity between the temporal lobe and AN.^20^ However, using the spectral GCA, we further clarified that delta frequency was the main outflow information from the thalamus to the seizure onset. To our knowledge, this study is the first to provide evidence demonstrating the selective involvement of the CM over the AN in frontal lobe seizures.

Successful treatment of epilepsy can be achieved by accurately stimulating the relevant thalamic nucleus involved in the seizure network. A human study showed that by stimulating the central lateral nucleus of the intralaminar thalamus during limbic seizures, cortical EEG signals were effectively restored, leading to improved arousal in individuals with limbic epilepsy.^31^ Another study further demonstrated that high-frequency stimulation (> 45 Hz) of the AN resulted in the desynchronization of ipsilateral hippocampal activity, successfully reducing seizure generation and propagation.^32^ Future studies will likely optimize stimulation parameters and elucidate the mechanisms underlying seizure modulation via CM or pulvinar stimulation.

This study has several limitations. The GCA typically requires signals to be stationary and linear,^33^ and it is worth noting that the EEG signals utilized in this study may not strictly adhere to these assumptions. We employed various strategies to mitigate the deviation from stationarity assumptions. These strategies included filtering techniques to eliminate power line noise^34^ and adopting windowing approaches to analyze shorter intervals, thereby enhancing the approximation to stationarity.^35^ Our primary objective in this study was to establish a connection between cortical seizure foci and the thalamus. However, we will consider using other sophisticated measures of causality^36, 37^ or non-linear regression approaches proposed in prior SEEG connectivity studies^20, 38^ to further verify our results. In terms of the study cohort, the sample size was small (n=10) and included individuals with heterogeneous epilepsy types. Notably, occipital lobe epilepsy was not included in this study. Additionally, our investigation solely focused on the AN and CM thalamic nuclei. As previous studies have emphasized the significance of the pulvinar nucleus in limbic and temporal lobe epilepsy, it would be of interest to extend the network analysis to incorporate the pulvinar nucleus. Furthermore, exploring connectivity changes at the end of seizure segments, as other research teams conducted,^9, 20^ would also be valuable.

In future studies, we plan to include more individuals with comprehensive thalamic coverage, as clinically indicated, including bithalamic or coverage of the pulvinar nucleus (in cases such as temporal, parietal, and occipital epilepsy). With the continuous progress in technology and increased awareness of neuromodulatory treatment options, it is anticipated that there will be a larger pool of candidates eligible for thalamic stimulation.^16, 39, 40^

## Data Availability

All data produced in the present study are available upon reasonable request to the authors.

## ACKNOWLEDGEMENTS

The authors have no conflict of interest to disclose. HN is supported by the National Institute of Neurological Disorders and Stroke (NINDS) K23NS128318, the Sudha Neelakantan & Venky Harinarayan Charitable Fund, the Elsie and Isaac Fogelman Endowment, and the UCLA Children’s Discovery and Innovation Institute (CDI) Junior Faculty Career Development Grant (#CDI-TTCF-07012021). AD is supported to research abroad by the Uehara Memorial Foundation and SENSHIN Medical Research Foundation. RS serves on scientific advisory boards and speakers bureaus and has received honoraria and funding for travel from Eisai, Greenwich Biosciences, UCB Pharma, Sunovion, Supernus, Lundbeck Pharma, Liva Nova, and West Therapeutics (advisory only); receives royalties from the publication of Pellock’s Pediatric Neurology (Demos Publishing, 2016) and Epilepsy: Mechanisms, Models, and Translational Perspectives (CRC Press, 2011). RS is also supported by the Sudha Neelakantan & Venky Harinarayan Charitable Fund.

We are indebted to Shaun A. Hussain, Joyce H. Matsumoto, Lekha M. Rao, Rajsekar R. Rajaraman, Maria Garcia Roca, Richard Le, Patrick Wilson, Cesar Dominguez, and Jimmy C Nguyen for their assistance in the study and sample acquisition.

## AUTHOR CONTRIBUTION

HN and WS contributed to the conception and design of the study. All authors contributed to the acquisition and analysis of data. SP, AD, BE, MM, SO, AF, RS, WS, and HN contributed to drafting/revising the text and preparing the figures.

---

**Supplementary Figure 1.**
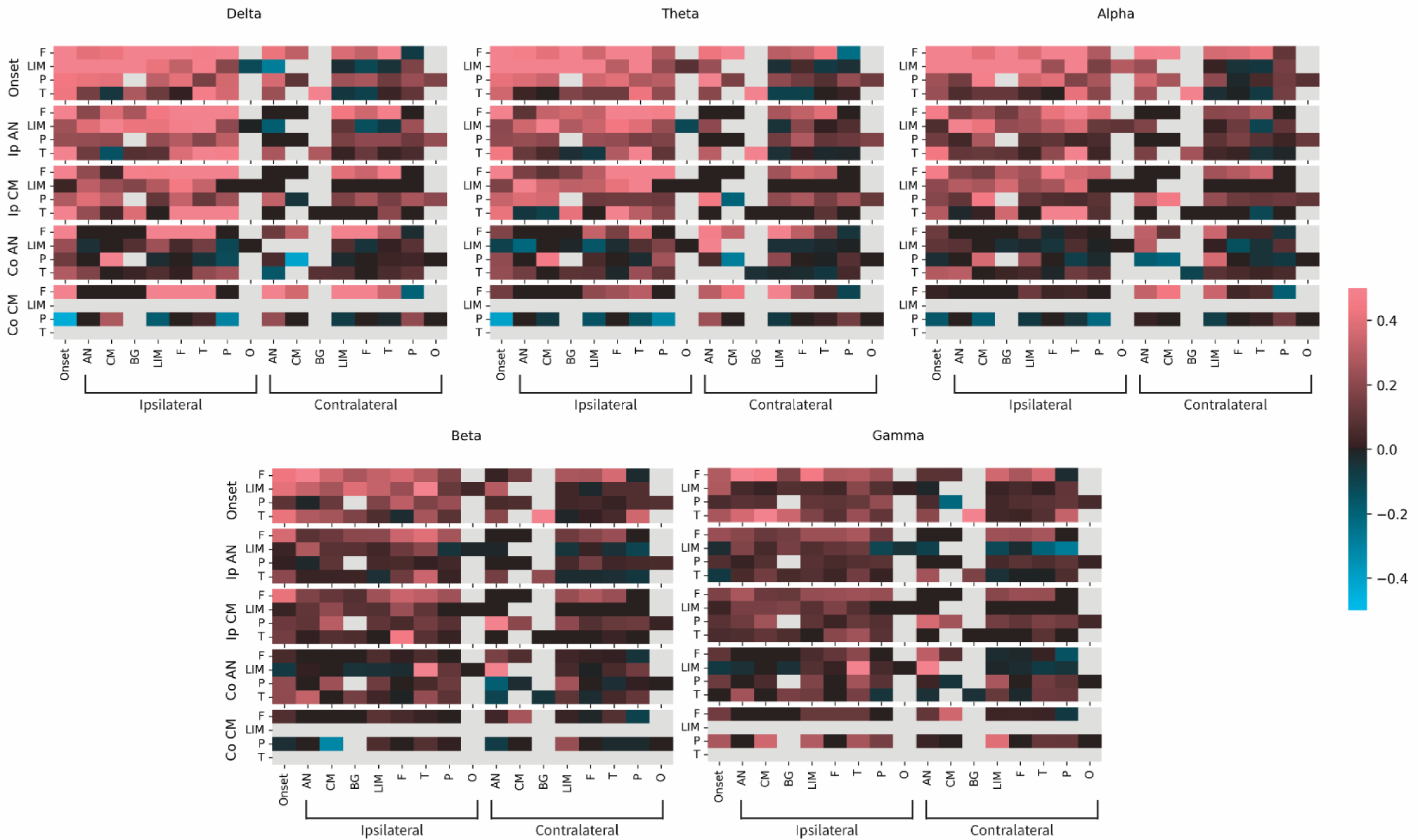
Percent change in spectral Granger causality inflow across lobes and frequency bands. Each heatmap demonstrates the percent change in spectral Granger causality from baseline (20-10 mins before seizure) to 0-10s after seizure for a particular frequency band. The heatmaps show the percent change in spectral Granger causality from a baseline period (20-10 minutes before seizure) to a specific time window after seizure onset (0-10 seconds). Each heatmap represents a particular frequency band. The rows in each heatmap represent the mean of seizures for a particular seizure onset lobe, and every grouping of 4 rows represents the inflow of causality from a particular set of electrodes (cortical onset and thalamus). Each column represents the electrode to which information is flowing. To illustrate how to interpret the figure, the cell in the bottom left of the delta heatmap represents the percent change in Granger causality between the onset region and the contralateral CM across temporal lobe seizures. (F = Frontal, LIM = Limbic, P = Parietal, T = Temporal, AN = Anterior Nucleus of the Thalamus, CM = Centromedian Nucleus of the thalamus, BG = Basal Ganglia, O = Occipital, Ip = ipsilateral, Co = contralateral).

**Supplementary Figure 2.**
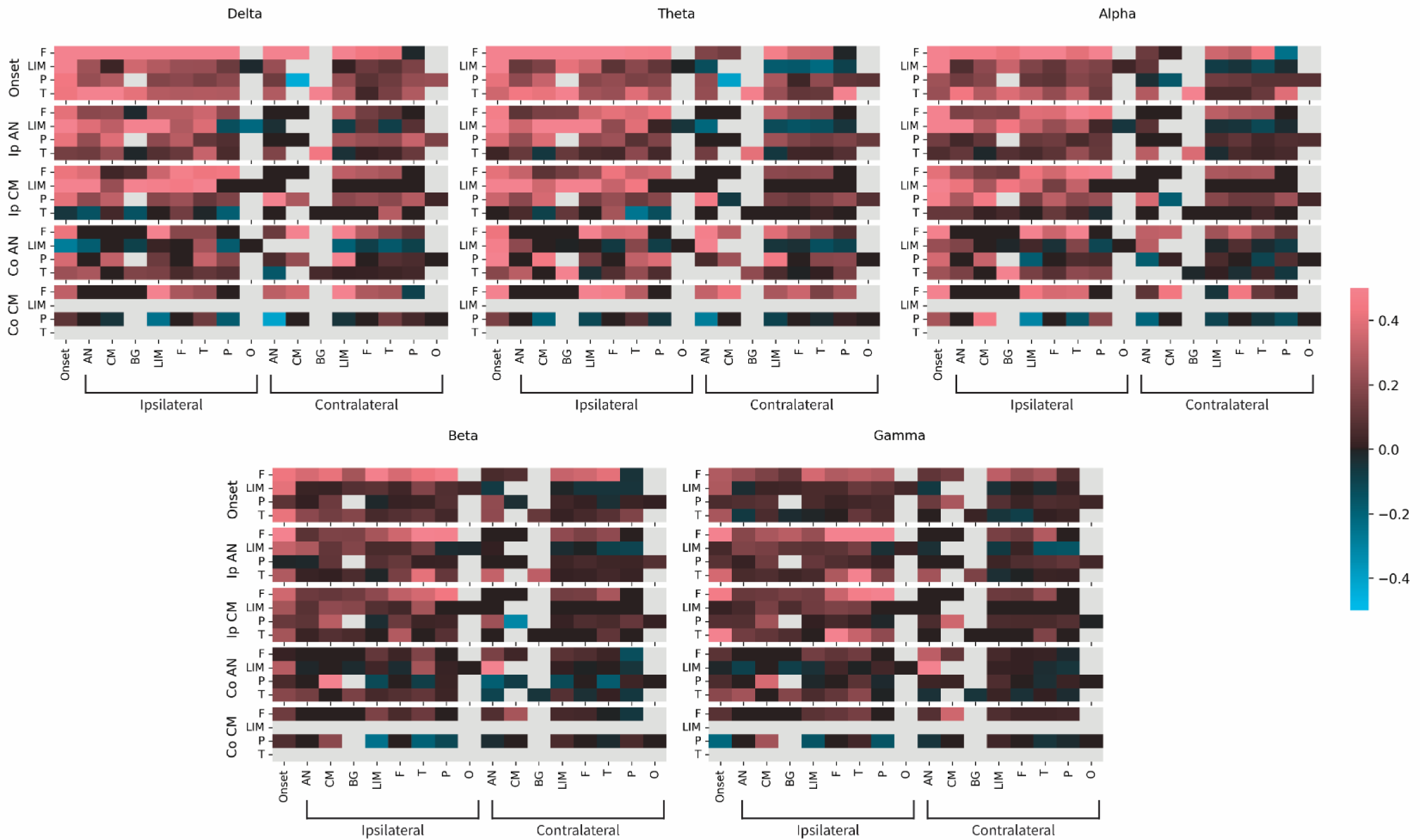
Percent change in spectral Granger causality outflow across lobes and frequency bands. This figure is similar to Suppl. Fig. 1, except each grouping of 4 rows represents the outflow of causality from the same set of electrodes.

